# Federated Learning for the pathogenicity annotation of genetic variants in multi-site clinical settings

**DOI:** 10.1101/2025.04.03.25325184

**Authors:** Nigreisy Montalvo, Francisco Requena, Emidio Capriotti, Antonio Rausell

## Abstract

Rare diseases collectively affect 5% of the population. However, fewer than 50% of rare disease patients receive a molecular diagnosis after whole genome sequencing. Supervised machine Learning is a valuable approach for the pathogenicity scoring of human genetic variants. However, existing methods are often trained on curated but limited central repositories, resulting in poor accuracy when tested on external cohorts. Yet, large collections of variants generated at hospitals and research institutions remain inaccessible to machine-learning purposes because of privacy and legal constraints. Federated learning (FL) algorithms have been recently developed enabling institutions to collaboratively train models without sharing their local datasets. Here, we present a proof-of-concept study evaluating the effectiveness of federated learning for the clinical classification of genetic variants. A comprehensive array of diverse FL strategies was assessed for coding and non-coding Single Nucleotide Variants as well as Copy Number Variants. Our results showed that federated models generally achieved comparable or superior performance to traditional centralized learning. In addition, federated models reached a robust generalization to independent sets with smaller data fractions as compared to their centralized model counterparts. Our findings support the adoption of FL to establish secure multi-institutional collaborations in human variant interpretation.

## Introduction

Rare diseases include a diverse range of more than 6172 clinical conditions that collectively affecting 4-5% of the population (1). Approximately 70% of rare diseases are considered to be of genetic origin (2). These are caused by high-impact germinal DNA defects including single-nucleotide variants (SNV) or large duplicated or deleted chromosomal regions referred to as Copy Number Variants (CNVs). Whole genome sequencing (WGS) has become a first-line genetic test for the diagnosis of rare diseases in the health system as it provides a comprehensive view of genomic alterations (3). Yet, the assessment of the approximately 5 million genetic variants typically found in a single individual genome is still challenging, and only an average of 40% of patients receive a molecular diagnosis (4,5).

Supervised machine learning (ML) has become a prominent bioinformatics strategy for the pathogenicity annotation of genetic variants, thanks to its ability to uncover complex patterns among genomic features (6,7). Here, models are trained on collections of genetic variants previously annotated as pathogenic or benign. Most state-of-the-art supervised ML scores for pathogenic variants prediction have been trained on curated genotype-phenotype databases, which encompass diverse sets of variant types, genes, diseases and phenotypic annotations (8). However, such reference databases are still limited in size and heterogeneity, and often translate in supervised ML scores that poorly generalize to newly encountered patients or disease cohorts (9). Such limitations are even more pronounced in the case of genetic variants affecting non-coding genomic regions, i.e. those outside protein-coding sequences, whose functional and clinical impacts are still largely uncharacterized (10,11).

Multi-institutional collaboration can increase genetic and clinical data size and diversity, and therefore, improve the generalization of ML models for variant assessment. However, regulatory policies, such as General Data Protection Regulation (GDPR) for European Union (12) and Health Insurance Portability and Accountability Act (HIPAA) in the United States of America (13), preclude genomic and clinical data sharing, as it represents threats to patient privacy. In genetic research, for example, data leaks can have harmful consequences for patients, such as genetic discrimination or data misuse (14,15).

Federated Learning (FL) has emerged as a promising solution for data-private collaboration in medicine and health (16–18). In contrast to collaborative data sharing (CDS), where institutions need to centralize their local datasets for model training, FL proposes keeping the data decentralized and learning a consensus model, by aggregating locally-computed updates. In the traditional implementation, the clients (e.g., hospitals or health-research institutions) are coordinated by a central server, which defines and maintains a global ML model. At each round of FL, the server sends a copy of the model parameters to the clients for local training, and aggregates the local updates to derive a new global model, which is used in the subsequent round (**Figure 1**). This process is repeated until a maximum number of rounds is achieved or a different stopping condition is met.

**Figure 1.**
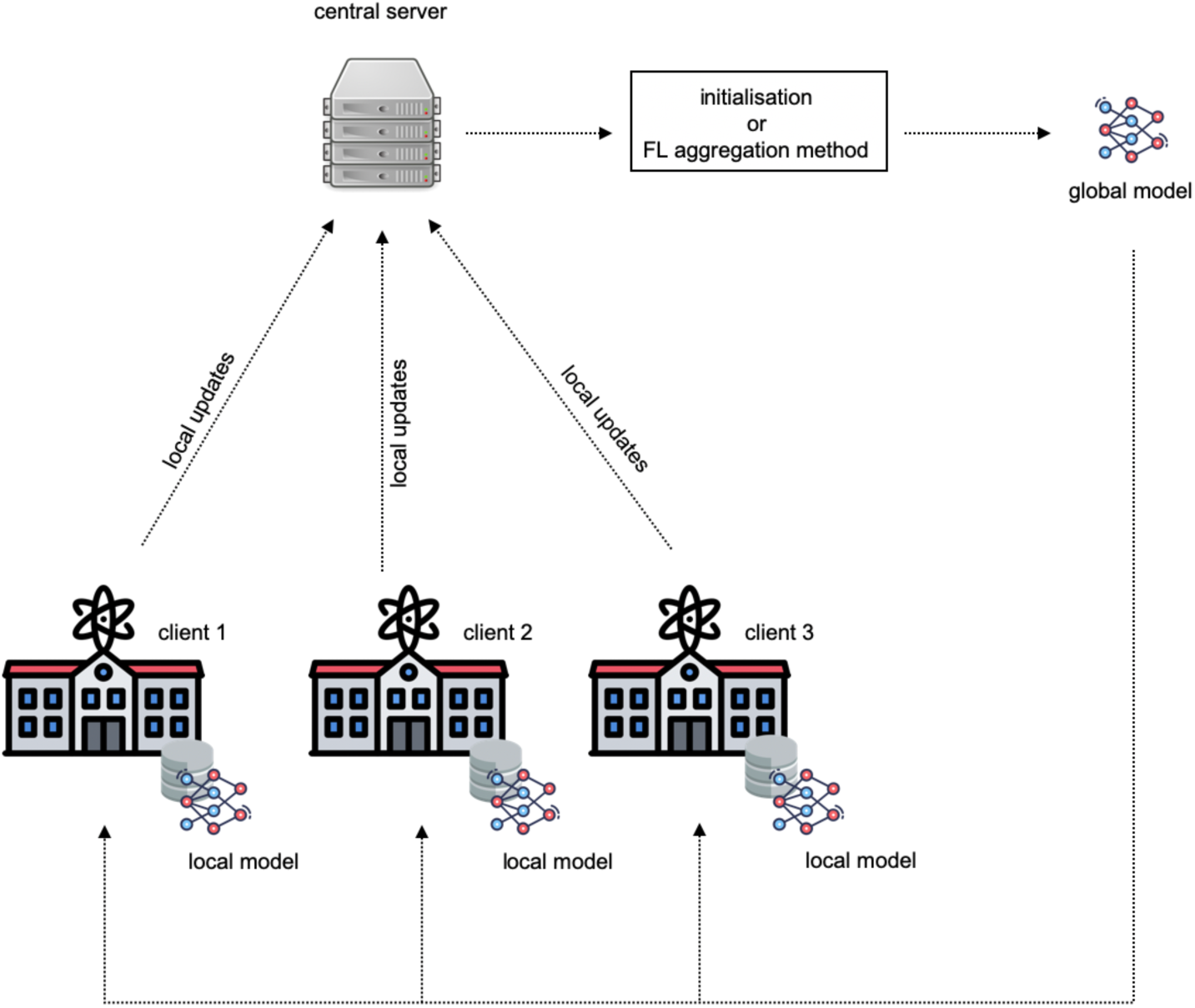
General overview of Federated Learning training. A central server orchestrates the collaborative training of a global machine learning model across the clients. Instead of raw data, only model parameters are exchanged between the server and clients.

In recent years, FL has proven effective for secure genomic data sharing. Nasirigerdeh et al. (19) presented sPLINK, a tool for the federated learning implementation of collaborative genome-wise association studies (GWAS). Experiments showed that sPLINK was robust against different sources of data heterogeneity, including the distribution of phenotypes and confounding factors. Raimondi et al. (20) proposed a FL solution for multi-site exome-based risk prediction of Crohn’s disease patients. The authors leveraged three public databases containing case and control Whole Exome Sequencing data to simulate a FL setting involving variable numbers of data owners. The results showed that the FL model improved the accuracy of models trained locally, even when the FL model was trained across 10 data owners who held very small datasets. More recently, Kolobkov et al. (21) proposed a simulated FL study for phenotype-from-genotype prediction and ancestry-from-genotype prediction on UK Biobank and the 1000 Genome Project. The authors showed that FL models were almost as accurate as the CDS model, and outperformed considerably the local models. To the best of our knowledge, however, studies evaluating the efficacy of FL for the pathogenicity annotation of genetic variants are currently lacking.

In this paper, we propose a proof-of-concept FL study for the pathogenicity annotation of genetic variants across independent institutions without raw data exposure. By leveraging the submitter information associated with each genetic variant in the publicly available ClinVar database (22), we mimicked three realistic multi-institutional collaborations for the clinical assessment of human genetic variants corresponding to three major types: coding SNVs, non-coding SNVs, and deletion CNVs. We then evaluated a comprehensive array of diverse FL strategies incorporating alternative network-based models, FL aggregation algorithms, local optimizers, client participation rates, FL server and client learning rates, collectively accounting for 1344 different settings (**Supplementary Table 1**). For each variant type, we systematically compared the performance of FL against that of CDS and single-institutional models. Model performance was assessed through cross-validation on the training set as well as on two additional independent test sets, allowing us to evaluate the generalization capabilities of the classifiers across the three case studies. Further experiments evaluated model robustness to client dropouts as well as model behavior under identically or non-identically distributed features. Our study showed that overall, the performance of federated models is generally superior to local models, and can reach comparable or superior results to CDS.

## Methods

### Genetic variants data collection

Coding and non-coding SNVs were extracted from ClinVar (version December 2020) as described in Capriotti and Fariselli 2023 (23). To adapt the dataset to a federated learning setting, we performed the following modifications: We first split variants into two non-overlapping subsets: 1) SNVs reported before January 1st 2020 (referred to as *SNVs-Before-2020-01*), and 2) SNVs reported after January 1^st^ (referred to as *SNVs-After-2020-01*). For coding SNVs, the multi-institutional training set was composed of SNVs from institutions having at least 1K coding SNVs in *SNVs-2020-01*, which resulted in 6 independent silos, with sizes ranging from 30,815 to 1,417 variants (**Supplementary Table 2**). Each institution subset was randomly downsampled in order to obtain a 1:1 balanced set of pathogenic and benign variants. In addition, we obtained 2 balanced independent test sets: a first set consisting on the remaining coding SNVs from *SNVs-2020-01* (10,378 variants) and a second set containing the coding SNVs from *SNVs-After-2020-01* (2,838 variants) respectively. In the case of non-coding SNVs, we obtained a multi-institutional training set formed by 8 institutions having at least 100 non-coding SNVs in *SNVs-Before-2020-01*, with sizes ranging from 2,288 to 101 (**Supplementary Table 3**). Similarly to coding SNVs, random downsampling was performed to obtain a 1:1 balanced set of pathogenic and benign variants and two independent test sets obtained containing 5,534 and 472 non-coding SNVs respectively.

In the case of CNVs, a high-confidence non-redundant set of pathogenic and benign deletion CNVs was obtained as described in Requena et al. (24). Pathogenic and likely pathogenic deletion CNVs were obtained from ClinVar (version October 2021). Benign CNVs were obtained from reference databases and matched by genomic length with the pathogenic CNVs in ratio 1:1 [3]. Similarly to coding and non-coding SNVs, we split the dataset into two non-overlapping subsets: 1) deletion CNVs reported before January 1st 2021 (referred to as *CNVs-Before-2021-01*), and 2) deletion CNVs reported after January 1^st^ (*CNVs-After-2021-01)*. For the purpose of this study, we considered benign deletion CNVs that had the same submission date as the corresponding pathogenic variant to which they were matched (see above). We derived a multi-institutional dataset by taking the deletion CNVs belonging to those institutions having at least 80 CNVs in *CNVs-Before-2021-01*, resulting in 8 independent silos, each with a 1:1 balanced set of pathogenic and benign variants, with sizes ranging from 6,936 to 82 variants (**Supplementary Table 4)**. We used the remaining deletion CNVs from *CNVs-Before-2021-01* to form the first independent test set, accounting for a total of 682 samples. A second independent test set was formed with the 96 samples from *CNVs-After-2021-01*. The largest silo in the obtained multi-institutional dataset held around 90% of the total data, while each of the remaining silos individually accounted for approximately 1-2%. In order to avoid bias during training, we decided to train centralized and federated models across the smaller clients and compare their performance with the model trained on the largest client.

### Variant features annotation

Both coding and non-coding SNVs were annotated with a total of 60 features following (23), including: i) 25 values representing the 5-nucleotide window sequence centered on the mutated position (5 times 5 possible nucleotides: A, C, G, T, N), ii) 10 values representing the conservation scores of 100-species (PhyloP100) and 470-species alignments (PhyloP470) of the five-nucleotide window sequence centered on the mutated position (25). In this study, we also annotated SNVs by using an additional 25 values mapping the conservation scores of 3-species (PhyloP3), 4-species (PhyloP4), 7-species (PhyloP7), 17-species (PhyloP17), and 20-species (PhyloP20) to the five nucleotide window positions.

CNVs were annotated with 38 features, grouped into gene-based and region-based features, as described in (24). CNV gene-based features involved genes for which at least one base pair overlapped with the CNV genomic coordinates, based on Ensembl Gene (version 103), and using the GRCh37.p13 human reference genome. Genes were annotated using the following features: i) the probability of loss-of-function intolerance of the gene (pLI version 2.1.1, (26)); ii) the loss-of-function observed/expected upper bound fraction (LOEUF score, version 2.1, (26)); iii) the probability of being tolerant to both heterozygous and homozygous loss-of-function variants (pNull, version 2.1.1, (26)); iv) the constrained coding region (CCR) score (27); v) the enhancer domain (EDS) score (28); vi) predictions of haploinsufficiency or triplosensitivity, based on a meta-analysis of rare CNVs from 753,994 individuals (29); vii) ohnolog genes, as reported in the OHNOLOGS database (version 2, (30)), considering only pairs labeled as strict (highly reliable); viii) genes encoding transcription factors, according to data from the FANTOM consortium; ix) fitness cost due to gene inactivation, based on a genome-wide CRISPR-based score (CRISPR score, (31)); x) involvement of the proteins encoded by the genes in a protein complex (as extracted from hu.MAP 2.0 (32), with only proteins labeled with extremely high confidence selected); xi) mean and minimum gene expression across 54 tissues, obtained from the median transcripts per million (TPM) expression levels for each gene, provided by the Genotype-Tissue Expression Project (GTEx, version 8, (33)), in which all GTEx tissues were considered; xii) mean PhastCons 46-way placental score (34), xiii) the CpG density of the promoter regions identified as 2 kb upstream and downstream from the transcription start site (TSS), defined as the first nucleotide of the transcript, according to previous work (35); and xiv) six network-based gene/protein features extracted from a protein-protein interaction network: degree, PageRank, and shortest path to proteins associated with haploinsufficient and triplosensitive genes respectively (24). Gene-based features were transformed into categorical variables (“0” or “1” coding for the absence or presence, respectively, of at least one gene with the corresponding categorical feature), or quantitative variables encoding the maximum or minimum of the corresponding feature across the genes mapping within the CNV, except for minimum expression, the shortest path to haploinsufficient genes, and the shortest path to triplosensitive genes, for which the minimum was applied.

CNV region-based features considered included: i) the percentage of the CNV covered by each of the following six types of regulatory regions: open chromatin regions, transcription factor binding sites (TFBS), promoters, promoter flanking regions, CTCF sites, and enhancers, identified on the H1 human embryonic stem cell line (H1-hESC) and obtained from the Ensembl Regulatory Build (version 2019-11-01, (36)); ii) the maximum recombination rate (37), CADD score (version 1.6, (38)) and GERP scores (39) across the CNV genomic interval, with the scores previously summarized by their maximum value within non-overlapping 100 base pairs (bp) sliding windows; iii) Maximum gene density across the 1 megabase (Mb) sliding windows overlapping the CNV genomic interval; iv) four features encoding the presence or absence of CNV overlap (i.e. at least a 1 bp overlap) with the following regions of biological interest: a) human accelerated regions (HARs) (40), b) lamina-associated domains (LADs) (41), c) ultra-conserved non-coding elements (UCNEs) (42)), and d) structural variant (SV) hotspot regions (43); and v) two features encoding the distance (in Mb) to the centromere and the closest telomere regions respectively; genomic coordinates were retrieved from the UCSC Genome Browser “Gap” track (44).

### Federated learning settings

In this study we focused on a cross-silo and horizontal FL setting. Cross-silo refers to a small number of participants, typically organizations, such as genetic testing companies and research institutions in our context. The organizations are always available for local training and can participate in each round of FL training. The term horizontal indicates that clients’ datasets share the same feature space, but hold different data samples. As illustrated in **Figure 1**, one round of FL encompasses the following steps: 1) The central server initializes a global ML model. In the experiment the models were initialized with random weights. 2) The central server then selects a subset of clients and broadcasts them a copy of the global model. Since we train neural network-based models, communicating the model refers to exchanging model weights. 3) The clients then use the local dataset for optimizing the received model through Stochastic Gradient Descent (SGD) for a predefined number of epochs. In the experiments, the number of local epochs was fixed to 10. 4) After completing the local training, the clients send the local updates (local models) to the central server. 5) The central server uses a FL aggregation method to combine the local updates into a new global model. Steps 2–5 are repeated for a predefined number of rounds, which was set to 200 in our experiments, or until a stopping condition is met.

Throughout the FL process, clients and central server kept the same neural network to ensure consistency in the learning process and allow for seamless aggregation of local updates from multiple clients. Different aggregation algorithms can be used by the server to integrate the local updates into a unified ML model. In this study we benchmarked the following FL aggregation algorithms: (i) FedProx (45) is a re-parametrization of Federated Averaging (FedAvg) (46), specifically designed to address statistical heterogeneity among clients. In FedAvg, the central server randomly selects a subset of clients for local optimization and aggregates their local models using a weighted averaging approach, where the weights are taken proportional to the clients’ training dataset sizes. FedProx extends FedAvg by introducing a regularization term to the local training loss, denoted as µ, which constrains the local updates to be close to the global model. Notably, FedAvg is a special case of FedProx, when μ = 0 and Stochastic Gradient Descent (SGD) is the local optimizer. (ii) FedAdagrad (47) is a federated version of the adaptive optimizer Adagrad (48). The central server adjusts the learning rate for each model parameter based on the historical gradient information, therefore improving model convergence on non-IID data. (iii) FedAdam (47) is an adaptation of the adaptive optimizer Adam (49) to the FL setting. The central server adjusts the learning rate for each model parameter by using the first moment (the mean) and the second moment (the uncentered variance) of the gradients. (iv) FedYogi (47) is a federated version of the adaptive optimizer Yogi (50), which proposes modifications to the update rule of the second moment in Adam to improve model stability in non-convex optimization scenarios. In the literature, FedAdagrad, FedAdam, and FedYogi are considered FedOpt variants.

### Neural network-based models

Two neural network-based models were evaluated in this study: (i) a prototypical and well known Multilayer Perceptron architecture (MLP) (51), and (ii) a Shallow Neural Decision Forest (sNDF) (52), for which provide here a short background for non-familiar readers. sNDF is a supervised ML model that combines elements of neural networks and decision trees. The main goal of sNDF is unifying the representation learning capabilities of deep neural networks with the divide-and-conquer mechanism of decision trees. To that end, sNDF implements first a deep convolutional neural network (CNN) for feature learning from raw input data, and then uses the learned representation for training a forest of stochastic and differentiable decision trees. Similar to Random Forest (53), each decision tree is trained using a subset of features. The final prediction of the forest is taken as the average of individual decision tree predictions. In this approach, the model takes the original feature vectors as input and feeds them into a forest of stochastic and differentiable decision trees, allowing for end-to-end training. More specifically, the original feature vectors are followed by a block of fully connected layers, with output units *fn* (52). Each *fn* is associated with a decision (split) node in a tree, with decision function *dn*(x) = *σ*(*fn*(x)), where *σ*(x) = (1 + e^−x^)^−1^ is the sigmoid function. The output of *dn* is interpreted as the probability of routing the sample to the left or right subtree. Once the sample reaches a leaf node *l*, the prediction of the tree is given by the class-distribution *πl*. Further details on the training of the model can be found in (52).

### Hyperparameter tuning and model training and evaluation

To prevent, by design, downstream contamination of training and testing variants, we split training variant datasets by chromosome as proposed in (23,24,54). More precisely, for each variant type and model configuration we trained a bundle of 23 classifiers, so that variants in a given chromosome were predicted using the classifier trained on variants from the remaining chromosomes. For example, we predicted CNVs in chromosome 2 by using a model trained on CNVs in chromosomes 1, 3, 4, etc. We refer to this approach as *leave-one-chromosome-out* training. We followed the guidelines in (55) for hyper-parameter tuning by grid-search. We first tuned the hyperparameters of centralized ML models for further comparison with the federated version. For centralized MLP and sNDF we tuned the hyperparameters related to model architecture, batch size, learning rate, and optimizer. Specifically, we trained a 3-layer MLP and considered the following hyperparameters for this model: 1) number of neurons in the hidden layer ∈ {3, 4, 5, 6, 7, 8, 9}, 2) batch size ∈ {4, 8, 16, 32}, 3) learning rate ∈ {0.1, 0.01, 0.001, 0.0001}, 4) optimizer: SGD (with a momentum of 0.9) and Adam (with β1 = 0.9 and β2 = 0.999). We fixed weight decay to 0.0001. For sNDF we considered the same hyper-parameter sets for batch size, learning rate, optimizer and weight decay. We also explored the following hyperparameters related to model architecture: 1) number of trees ∈ {3, 5, 10, 15}, depth of the trees ∈ {3, 6, 9}, feature rate ∈ {0.5, 0.6, 0.7, 0.8, 0.9} (referring to the rate of original features used for training each individual tree in the forest). We used the best hyperparameters found per model (based on area under ROC curve) for training single-site models as well; and reused the architecture, batch size, and weight decay of the best centralized models for training the federated versions.

We optimized the FL-related hyperparameters that, for practicality, we divided into general, and FL aggregation method-specific. Among general hyperparameters we considered: 1) local learning rate ∈ {0.1, 0.01, 0.001, 0.0001}, 2) local optimizer: SGD (with momentum 0.9) and Adam (with β1 = 0.9 and β2 = 0.999), and 3) client rate ∈ {0.5, 1.} (referring to the number of clients selected randomly for local training). For FedProx we also consider proximal term μ ∈ {0.5, 0.1, 0.01, 0.001, 0.0001, 0}. For FedAdam, FedAdagrad, and FedYogi we also considered: 1) server learning rate ∈ {1, 0.1, 0.01, 0.001}, 2) τ ∈ {0.0001, 0.01, 0.1}, representing the adaptivity parameter. For FedAdam and FedYogi we fixed β1 = 0.9 and β2 = 0.999. In the case of the training of federated MLP we also considered the inclusion (or exclusion) of local batch normalization layers. **Supplementary Table 5** outlines the parameters retained for CDS and FL models for the pathogenicity annotation of coding SNVs, non-coding SNVs, and CNVs. Hyperparameters of CDS and FL models were tuned with 10-fold collaborative cross-validation (CCV) on the entire training data. As outlined in (56), CCV proposes coordinating cross-validation across participants in the federation, so that each institution splits its local dataset into training and validation sets. Therefore, the final validation set is the aggregation of each institutional validation set. To ensure fairness in the experiments, we used the same splits for comparing CDS and FL approaches. We performed CCV with 3 different seeds for model weight initialization. All models were trained to 200 epochs, with early stopping if the best model did not improve for 20 consecutive epochs. After finding the best parameters, we trained centralized (CDS), single-institutional (local), and FL models on the entire training set, and evaluated them in the two held-out independent test sets. In this case, we repeated the experiments with 30 different seeds for model weight initialization to obtain a more accurate estimation of model performance.

### Generation of identically-distributed settings for evaluation in federated learning models

The study of the potential impact of identical or non-identical feature distributions in the FL process was carried out here following a strategy described in (55), where identically distributed (IID) partitions were generated from the original training set. In order to do so, we took the complete training set and randomly distributed the variants across clients while respecting their original sample size. Then we trained the FL models on such new data partitions for 30 seeds for model initialization. We repeated the random data partition 100 times to obtain a histogram with the distribution of median AUC ROC values obtained for each training set.

### Implementation

We implemented all the experiments using Flower (57), a Python library that enables the federation of several ML frameworks, including PyTorch. The experiments were performed on a high-performance computing (HPC) cluster, consisting of 18 computed nodes, each equipped with between 16 and 24 CPUs.

## Results

### Realistic scenarios of collaborative training for genetic variant assessment across independent institutions

In this work we evaluated the performance in the classification of human genetic variants as pathogenic or non-pathogenic of a diverse set of supervised learning approaches trained in a Federated Learning (FL) manner across multiple institutions, and compared it to the collaborative data sharing (CDS) and individual-institution model counterparts **(Methods**). To that aim, we took advantage of the publicly-available ClinVar database, which reports the pathogenic classification of genetic variants primarily submitted by genetic testing laboratories and research institutions. ClinVar database allows the association of each variant to its submitter institution, which is leveraged in this study for mimicking a realistic multi-institutional collaboration with or without data sharing. Three major types of genetic variants were independently evaluated: coding SNVs, non-coding SNVs and CNVs. For each variant type, we defined a multi-institutional dataset for training FL and CDS models, as well as two independent test sets for assessing the performance of the classifiers (**Methods** and **Supplementary Table 1**). **Figure 2** shows the distribution of the training data across the institutions considered for each case study. Exact size of silos in the multi-institutional training set and in two independent test sets for each case studies are provided in **Supplementary Tables 2 - 4**.

**Figure 2.**
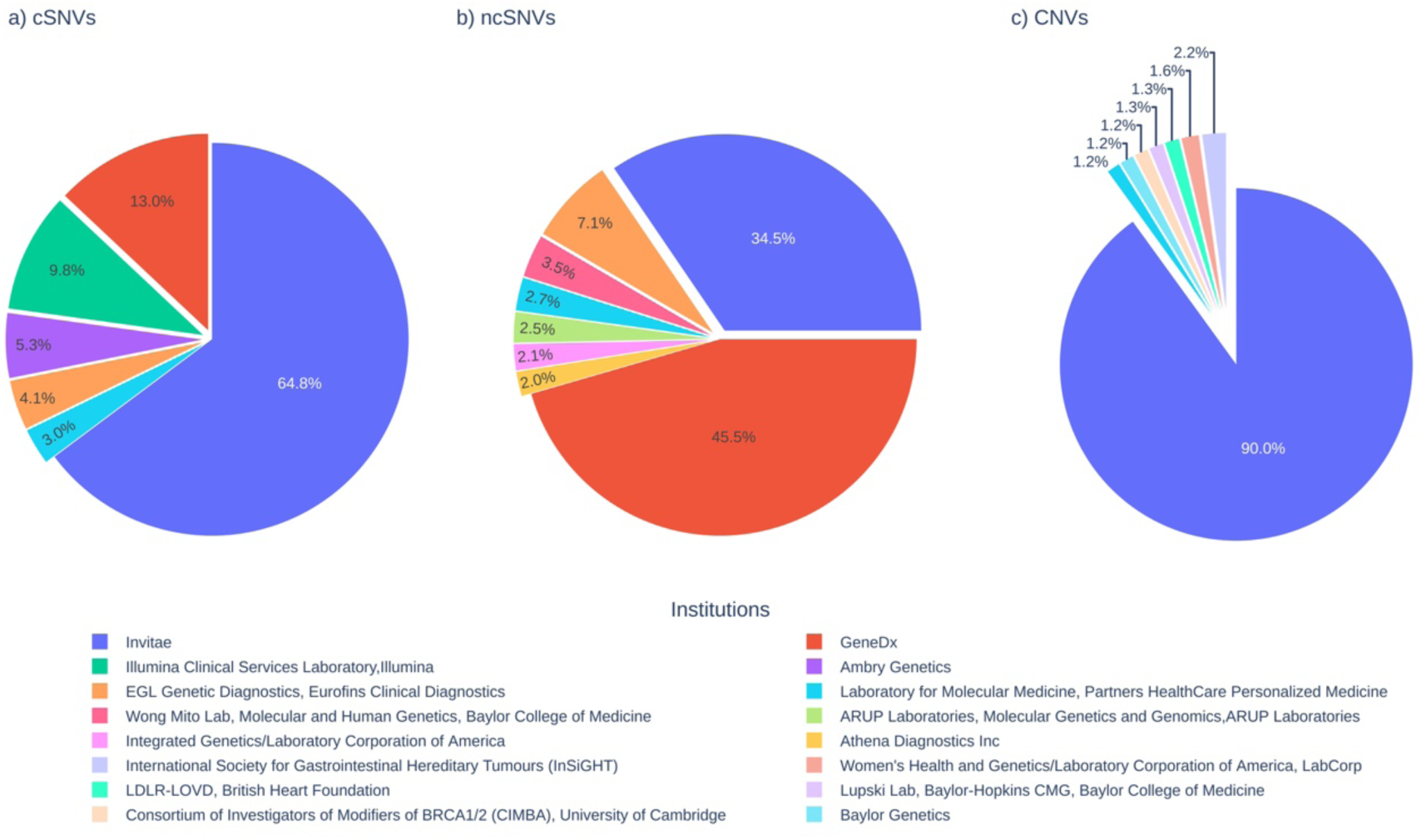
Distribution of the training data across institutions considered in the experimentation. Distributions are shown for a) coding Single Nucleotide Variants (cSNVs), b) non-coding Single Nucleotide Variants (ncSNVs), and c) deletion Copy Number Variants (CNVs).

The uneven distribution of variants observed across the participant institutions naturally provided different FL scenarios. In the case of coding and non-coding SNVs, the fraction of contributed instances showed a large heterogeneity across institutions, with contributors ranging from a maximum of 64.8% and 45.5% to a minimum of 3% and 2%, respectively (**Figure 2**). In contrast, CNVs presented a high concentration of variants in a single institution (90%) as compared to a panel of 7 small contributors whose contribution ranged between 1.2% and 2.2%. Thus, the first two cases were well adapted for a comparative analysis between the CDS and FL models involving all institutions. However, the third case appeared better suited for an evaluation comparing the performances of the models trained on the largest contributor against those resulting from the collaboration among the smaller ones, either in a CDS or a FL manner.

### Performance assessment of alternative federated learning model architectures

For each of the case studies described above we implemented two neural network-based models: a simple Multilayer Perceptron (MLP) as the first model (51) using ReLU activation functions, and a machine learning model combining neural networks and decision trees, named Shallow Neural Decision Forest (sNDF) (52) (**Methods**). For either the MLP or the sNDF models, we explored alternative FL strategies depending on the type of aggregation algorithm and the client rate used. The aggregation algorithm, defined as the mechanism used by the central server to combine the local updates into a unified ML model (**Figure 1**), plays a key role in FL, as it directly impacts model accuracy and convergence. Consequently, we benchmarked four FL aggregation algorithms, namely FedProx, FedAdagrad, FedAdam and FedYogi (45,47). Further details about the specificities of each method are provided in **Methods**. Second, we explored different values for FL client rate – the percentage of clients considered in a given round for local training by randomly selecting the clients. In addition, we evaluated the impact of the inclusion or exclusion of local batch normalization layers during the training of federated MLP, since several studies have proposed its use for improving FL model performance under non-independent and non-identically distributed (non-IID) data conditions (58,59).

We assessed model performance through cross-validation on the training set as well as on two additional independent test sets, allowing us to evaluate the generalization capabilities of the classifiers (**Methods**). The two independent test sets were designed to consider two complementary aspects: 1) variants belonging to cohorts from sites not having participated in the FL process, and 2) variants reported later than those variants used for FL training (**Supplementary Table 6**). **T**he different FL settings evaluated are summarized in **Supplementary Table 1**. Additional details on the collection of datasets, feature annotations, ML models used for training, and FL aggregation strategies are provided in **Methods**.

**Supplementary Figure 1** displays the area under the receiver operating characteristic (AUC ROC) curve obtained through cross-validation on the training set for the three types of genetic variants considered and across the different settings evaluated. **Supplementary Table 5** outlines the best hyperparameters retained for each evaluation setting. The results showed that FedProx generally outperformed the alternative FL aggregation strategies evaluated. In addition, we observed that a FL client rate of 50% generally led to improved performance across all FL aggregation algorithms. Finally, for the MLP models, the inclusion of a batch normalization layer was generally detrimental. Overall, the two alternative learning strategies, MLP and sNDF, were similarly competitive when using the optimal FL settings, i.e., FedProx optimization, a 50% client rate, and no batch normalization layer. Results observed in the two independent tests confirmed these general trends (**Supplementary Table 6**).

### Comparative performance evaluation among local-client, centralized, and federated models on two independent test sets

The assessment of the federated learning models through cross-validation on the training set allowed us to identify the optimal FL parameters for each type of genetic variant and learning method considered (MLP and sNDF), in terms of aggregation strategy, client rate, and batch normalization in the case of MLP (**Methods** and **Supplementary Table 5**). We then compared the performance of such optimal FL models against models obtained either from individual-client models or from centralized model counterparts, i.e., trained on aggregated client data. The AUC ROC values obtained on the two independent test sets for the three types of genetic variants considered are represented for the MLP and sNDF models, respectively, in **Figures 3** and **Supplementary Figure 2,** with complete details reported in **Supplementary Tables 7–9**.

**Figure 3.**
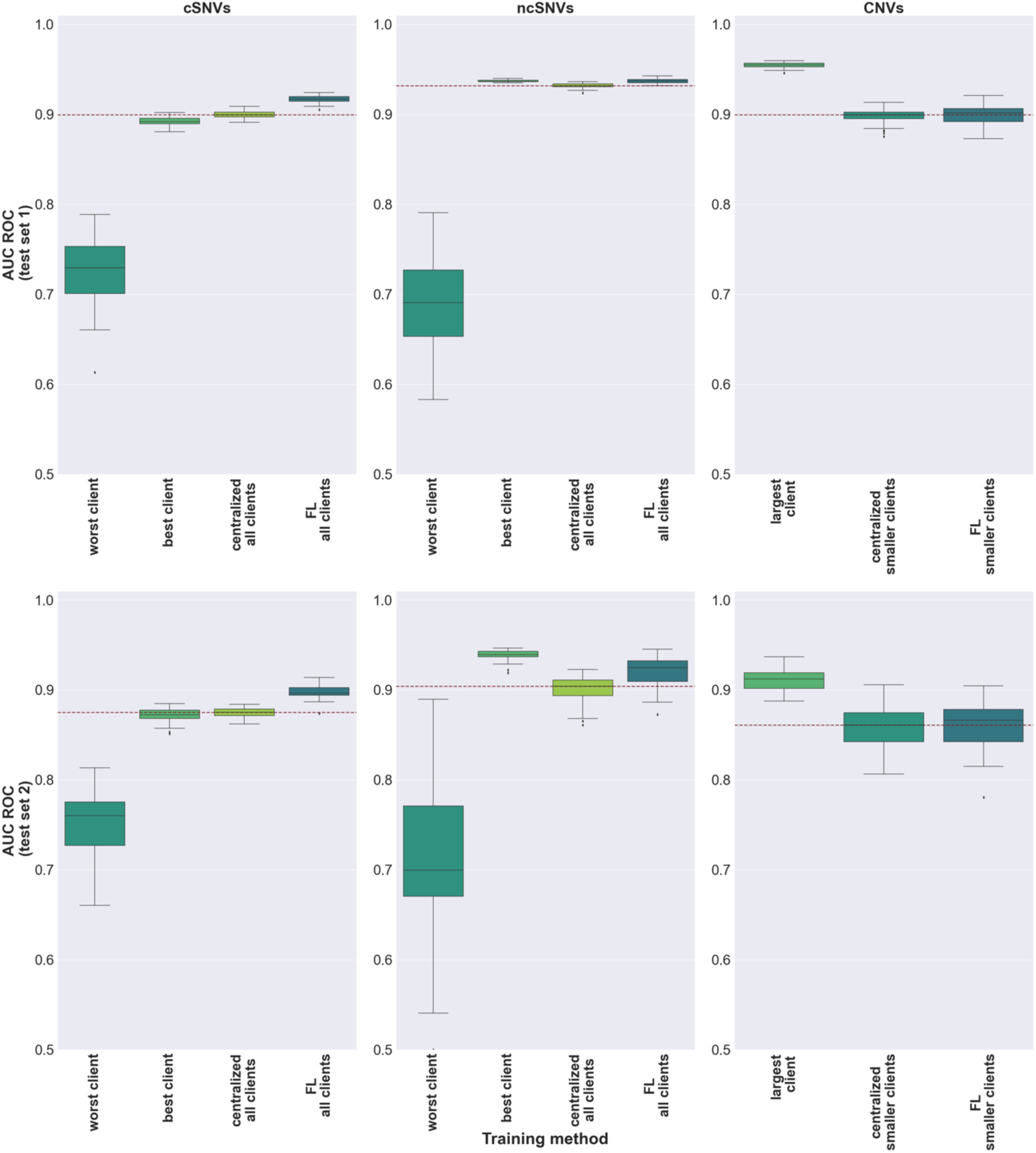
Performance of local, centralized, and federated MLP models on two independent test sets. Top and bottom panels show, respectively, the results on the first and second independent sets (Methods). In the case of coding SNVs and non-coding SNVs, the performance of the worst and best individual client models as well as the pooled-clients models are displayed. The performances of the remaining client models are reported in **Supplementary Tables 7–9**. In the case of CNVs, the performance of the largest client was compared against the centralized and federated learning models of the smaller clients. Boxplots in the panels represent the distribution of AUC ROC values obtained upon 30 different random seeds for model weight initialization. To ease comparison across models, a dotted red line represents the median value obtained for the centralized models.

In the case of coding and non-coding SNVs, the results showed that the federated learning models were consistently either comparable or superior to the centralized model counterparts. FL models were particularly remarkable in the case of MLP (**Figure 3**), where they significantly outperformed the centralized models in the two independent sets evaluated both for coding and non-coding SNVs (Wilcoxon rank sum test p-value <1.5e-05, **Supplementary Table 10**). Individual client models however showed a large heterogeneity, illustrating the risks of non-collaborative settings. Thus, while some reached competitive performance, others led to significantly inferior results (**Supplementary Tables 7–8**).

As previously introduced, in the case of CNVs the performance of the largest client model was compared against the centralized and FL models across the smaller clients. Here, the largest-client ranked first across all scenarios evaluated, as expected from the sharp difference in the training set size (**Figure 2**). However, despite this limitation, the cooperation across small clients through FL led to competitive results which, in the case of MLP, were not significantly different from their centralized versions (**Figure 4**). These results showed the beneficial impact of collaboratively training supervised models across different scenarios for the classification of human genetic variants while respecting data privacy constraints.

**Figure 4.**
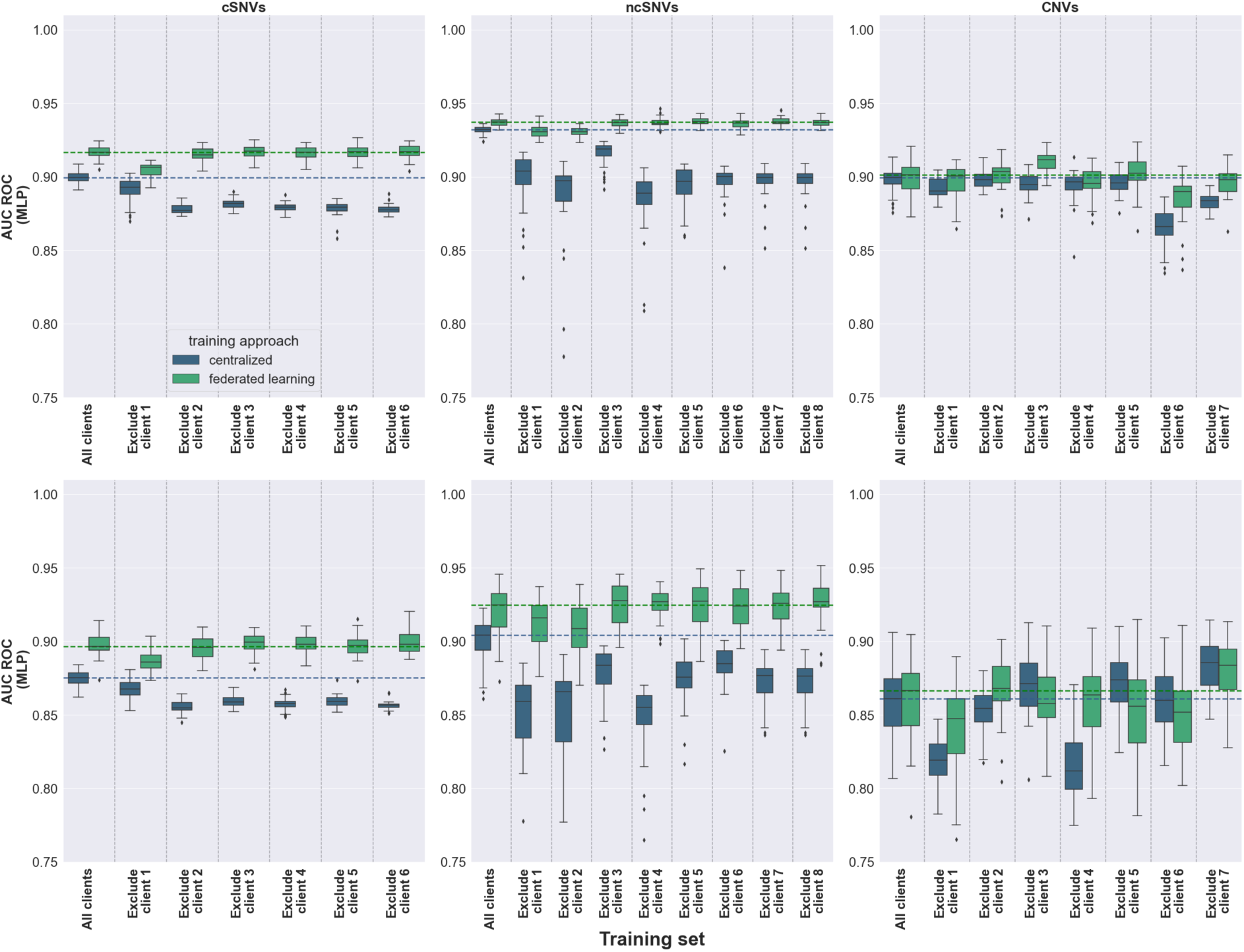
Performance of centralized and federated MLP models upon client dropouts on two independent test sets. Top and bottom panels show, respectively, the results obtained on the first and second independent test (Methods). Boxplots in the panels represent the distribution of AUC ROC values obtained upon 30 different random seeds for model weight initialization. Centralized and FL models are colored in blue and green, respectively. Each panel represents the AUC ROC values obtained considering all clients, as well as excluding one client at a time. To ease comparison across models, dotted lines represent the median values obtained for the centralized (blue) and federated (green) models considering all clients.

### Measuring the robustness of centralized and federated models against client dropouts

We then evaluated to what extent the performance of centralized and federated models would have been affected in the eventual case that a given client had not participated in the collaborative process. **Figures 4** and **Supplementary Figure 3** allow comparison of such events on the two learning algorithms evaluated, MLP and sNDF, respectively, for the two independent sets evaluated. In the case of coding and non-coding SNVs, the results showed that the federated learning models were consistently more resilient to client dropouts than the centralized model counterparts (**Supplementary Tables 11 – 12**). FL models were particularly robust in the case of MLP, where they were not significantly affected for most of the client drops, and in sharp contrast to the performance drops observed in the centralized settings (**Figure 4** and **Supplementary Figure 3**). In the case of CNVs, less consistent trends were observed upon client dropouts, reflecting stochastic sampling effects as a result of the small sample size on both the centralized and federated settings. Thus, while MLP models were generally robust to client dropouts in both the centralized and FL settings, sNDF showed less resilient to the absence of clients. Considered together, our results suggest that federated learning models need comparatively smaller training datasets than their centralized model counterparts in order to generalize adequately to independent datasets.

### Assessing the similarity of centralized and federated models

In the previous sections we showed that centralized and federated models led to overall comparable performances across diverse settings and scenarios, and this in spite of their different training process. Here we further inquired whether such results reflected a convergence of both approaches into largely redundant models, not only in terms of global accuracy, but also in terms of their classification outcomes for each specific genetic variant evaluated. To address this question, we evaluated the Spearman rank correlation between the pathogenicity scores for each genetic variant produced by the centralized and the federated model, for each evaluated setting (**Supplementary Table 13**). The observed rank correlation values ranged between 0.92 and 0.97, with p-values < 2.16e-132 for comparisons involving coding and non-coding SNVs, suggesting that models learned in a centralized and federated learning manner were largely equivalent for individual variant assessment. In the case of CNVs, the similarities between the two approaches were also statistically significant across all settings (p-values < 1.36e-25) yet with lower rank correlation values, ranging between 0.82 and 0.87. Such values suggest a larger divergence between the centralized and federated approaches as compared to the SNV scenarios, probably reflecting stochastic sampling variance across clients in the federated learning process due to the smaller clients’ sample size.

### Analyzing the influence of genetic variant feature distribution on federated learning performance

Motivated by the previous results, we further investigated whether the comparable or superior performances observed in FL as compared to centralized models were driven by underlying identical or non-identical distributions of genetic variant features across the clients participating in the training process. To this end, we first inspected the distribution of variants in the training set across an unsupervised low-dimensional representation obtained from the variants feature matrices (**Supplementary Figure 4**, **Methods**). In the case of coding and non-coding SNVs, variants were homogeneously distributed across the UMAP space both in terms of their clinical label (pathogenic or benign) and of their institutional submitter. In the case of CNVs, however, pathogenic variants formed distinctive clusters, which generally reflected their client of origin (**Supplementary Figure 4**). Those observations suggest that CNV variants, but not coding and non-coding SNVs, might have a non-identical feature distribution across participant Institutions involved in the FL experiments.

To further investigate this possibility, we then generated a pool of 100 synthetic random partitions of the initial training set across the same number of virtual clients, while respecting their original sample size, and trained FL models using the optimal hyperparameters on the generated partitions (**Methods**). For execution time limitations, these experiments were conducted exclusively on the MLP model. **Figure 5** shows the histograms representing the distribution of median AUC ROC values achieved by the FL models trained in the 100 newly generated training sets, and evaluated on the two independent test sets. In the case of coding and non-coding SNVs, the results showed that the actual median AUC ROC values obtained by the original model on the two independent test sets were not significantly different than those obtained by models where variants were randomly distributed across clients (z-scores ranging from −1.05 to 0.80, with normal p-values > 0.05). These observations reflect that both coding and non-coding Single Nucleotide variants may not exhibit strong non-identical feature distributions. Conversely, for CNVs, the original AUC ROC value was significantly lower than the random distribution in the case of the first held-out test set (z-score = −3.74, normal p-value = 0.74), while no significant difference was observed for the second test set. We hypothesize that, in this case, the limited sample size of the second test set (96 samples) might have reduced the statistical power to detect differences in performance. Nonetheless, the results observed on CNVs are in line with the observations from the low-dimensional representations, both reflecting that the actual CNV data partition across clients presented non-identically distributed features, probably accentuated by the small clients’ data size.

**Figure 5.**
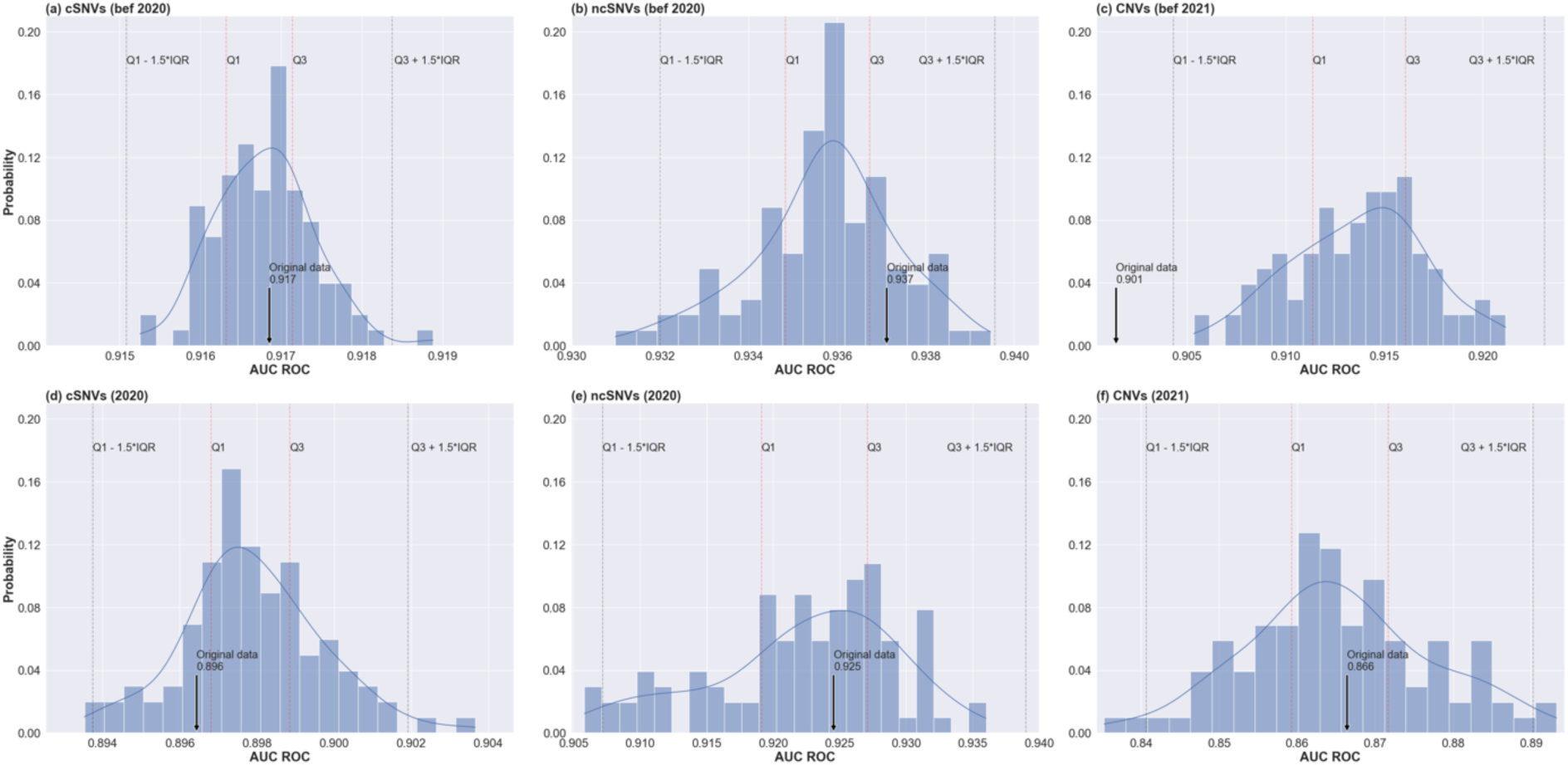
Distribution of median AUC ROC values obtained by the FL models trained on 100 randomly generated training sets. Top and bottom panels illustrate the histogram of the FL models evaluated, respectively, on the first and second independent test set for coding SNVs (left), non-coding SNVs (middle), and CNVs (right). For each histogram we delimit: the interquartile range (IQR = Q3 − Q1), representing the range within which the middle 50% of the data lies; the lower bound (Q1 − 1.5 × IQR) and upper bound (Q3 + 1.5 × IQR), for the identification of outliers; and the median area under the ROC curve (referred to as the original data) achieved by the FL model trained on the original data split.

The previous results suggest that the advantages observed for federated models throughout this work in terms of superior and more robust performance, would be driven by the iterative learning process across disjoint and partly represented datasets (with client rates < 100%) rather than by non-identical feature distribution across clients. As a corollary of this result, it may be proposed that, even when a large aggregated training dataset is available, mimicking a virtual federated learning through random splits across synthetic clients could lead to performance improvements.

## Discussion

In this work we carried out a proof-of-concept study evaluating whether Federated Learning (FL) might be an effective collaborative machine learning strategy for the pathogenicity annotation of human genetic variants in a clinical genomics context. To this end, we leveraged the publicly available database ClinVar to mimic realistic multi-institutional collaborations for the training of supervised ML models, with and without data sharing. Our experiments showed that, in most cases, FL achieved competitive or superior performance compared to collaborative data sharing (CDS) approaches, while also outperforming single-institutional models for the majority of participants. We also demonstrated that FL generally exhibited greater robustness to the removal of a participant’s data from the training set compared to CDS approaches. Such results suggest that FL needs relatively smaller datasets than CDS approaches to generalize adequately to unseen datasets. Our study thus supports FL as a beneficial approach for training supervised ML models for the clinical classification of genetic variants across multiple institutions, while respecting data privacy constraints.

Notably, under the settings based on a Multilayer Perceptron, FL outperformed CDS approaches in the clinical assessment of coding as well as of non-coding SNVs, despite the underlying identical distributions of genetic variant features across the clients participating in the training process. Such a result may initially seem counterintuitive, considering that a CDS approach benefits from complete data access and centralized optimization. However, our findings align with previous studies observing a similar trend in biomedical applications. For example, Linardos et al. (56) observed that a FL model for cardiac magnetic resonance imaging (MRI) diagnosis outperformed its centralized counterpart. The authors hypothesized that this behavior could be attributed to the model averaging process in each round of FL, which may have had a stabilizing effect, leading to improved performance across different model initializations. Ogier du Terrail et al. (55) presented a FL benchmark using seven multi-institutional datasets, simulating diverse realistic healthcare settings covering different tasks and modalities. Authors found that FL outperformed its centralized model counterpart when the ML models were linear and when the training set was low-dimensional, tabular data. Based on these studies, we speculate that FL outperformed the CDS approach in our case due to model averaging and to the characteristics of the training set, which consisted of low-dimensional tabular data. Interestingly, the performance gap between FL and CDS was more pronounced for MLP, a simpler model with fewer parameters, as compared to sNDF. This suggests that federated optimization may become more challenging as model complexity and number of model parameters increases.

The results obtained through cross-validation (**Figure 3**) revealed additional interesting findings. First, the introduction of batch normalization layers to local MLP models had a negative impact, damaging the performance of all FL aggregation strategies evaluated. While some studies suggested that the use of local batch normalization layers can enhance the performance of FL under conditions of data heterogeneity (58,59), other studies point to potential drawbacks, especially when there is a great mismatch between local and global model statistics (60). It is worth noting that these studies focused, however, on computer vision applications often involving deep learning architectures. In the context of our study, our results suggest that incorporating batch normalization layers into shallow models, such as a 3-layer MLP, can be counterproductive. Second, using only 50% of clients for local training, randomly selected in each round, generally resulted in better performance compared to using all clients. This observation suggests that, by training on a different subset of clients in each round, the FL model may avoid overfitting to specific clients’ data, improving its generalization capabilities. Finally, a corollary of our work is that, even in scenarios with full data access to large centralized genetic variant collections, mimicking FL training across virtual clients with randomly generated data splits can lead to model improvements over their centralized counterparts.

Supervised learning approaches for the clinical assessment of genetic variants have extensively relied on centralized curated repositories such as ClinVar (22) and Decipher (61), among others. While these are valuable resources for the community, they necessarily exclude sensitive information from the carrier individuals, including complete genomic data, clinical history and relevant phenotypic information. Thus, as our proof-of-concept study is based on such type of centralized repositories, we could only evaluate the beneficial aspects of FL models for variant classification that rely exclusively on genomic features of the variants, without considering patients’ clinical signs. Yet, our study serves as an incentive for the implementation of a FL approach in real-word settings that could benefit from the incorporation of additional data modalities also requiring data privacy, such as electronic health records and medical imagine.

In our experiments we assumed that clients and server are honest and trusted, which means that no party is attempting to disrupt or manipulate the FL training process. However, a real implementation of FL would need to take into consideration further privacy-preserving guarantees against potential malicious attacks, such as attempts to reconstruct the original data from model updates (62). Homomorphic encryption (63) and Differential Privacy (64) can be used to secure parameter exchange in FL. Homomorphic encryption allows computation to be performed on encrypted data, allowing the clients to encrypt the model updates before sending them to the server. The central server then aggregates the encrypted local updates, returning an encrypted global model to the clients for decryption. Although Homomorphic encryption offers strong privacy protections, current implementations are computationally expensive, slowing down the FL training process. Differential Privacy, on the other hand, adds noise to local updates before sending them to the server for model aggregation. However, it is crucial to carefully choose the amount of noise to be added, since excessive noise can degrade model performance, while insufficient noise may not provide sufficient privacy protection.

Our work extends to genetic variant assessment the scope of biomedical applications for which Federated Learning has proven to be an effective strategy for implementing collaborative machine learning approaches under data privacy constraints. These findings represent a major novelty in the field of clinical genomics and we expect them to encourage the adoption of FL to establish secure multi-institutional collaborations for human variant interpretation. This, in turn, would lead to more robust ML models that benefit not only from larger datasets but, most importantly, from more diverse datasets covering a wider range of genetic conditions, genetic backgrounds, and clinical manifestations.

## Declarations

### Ethics approval and consent to participate

Not applicable

### Consent for publication

Not applicable

### Availability of data and materials

All data required to reproduce the results presented in this manuscript, including figures and tables, will be made available under the GNU General Public License v3 upon publication. The data will be accessible via a *Figshare* repository.

All code required to reproduce the results presented in this manuscript, including figures and tables, will be made available under the GNU General Public License v3 upon publication. The code will be accessible via a *GitHub* repository at https://github.com/RausellLab/FedLearnVar

### Author Contributions

Conceived and designed the experiments: NM, AR. Performed the experiments: NM. Analyzed the data: NM, AR. Contributed materials/analysis tools: NM, FR, EC, AR. Wrote the paper: NM, EC, AR.

## Data Availability

All data required to reproduce the results presented in this manuscript, including figures and tables, will be made available under the GNU General Public License v3 upon publication. The data will be accessible via a Figshare repository.
All code required to reproduce the results presented in this manuscript, including figures and tables, will be made available under the GNU General Public License v3 upon publication. The code will be accessible via a GitHub repository at https://github.com/RausellLab/FedLearnVar

## Supplementary data

### Supplementary Figures

**Supplementary Figure 1:**
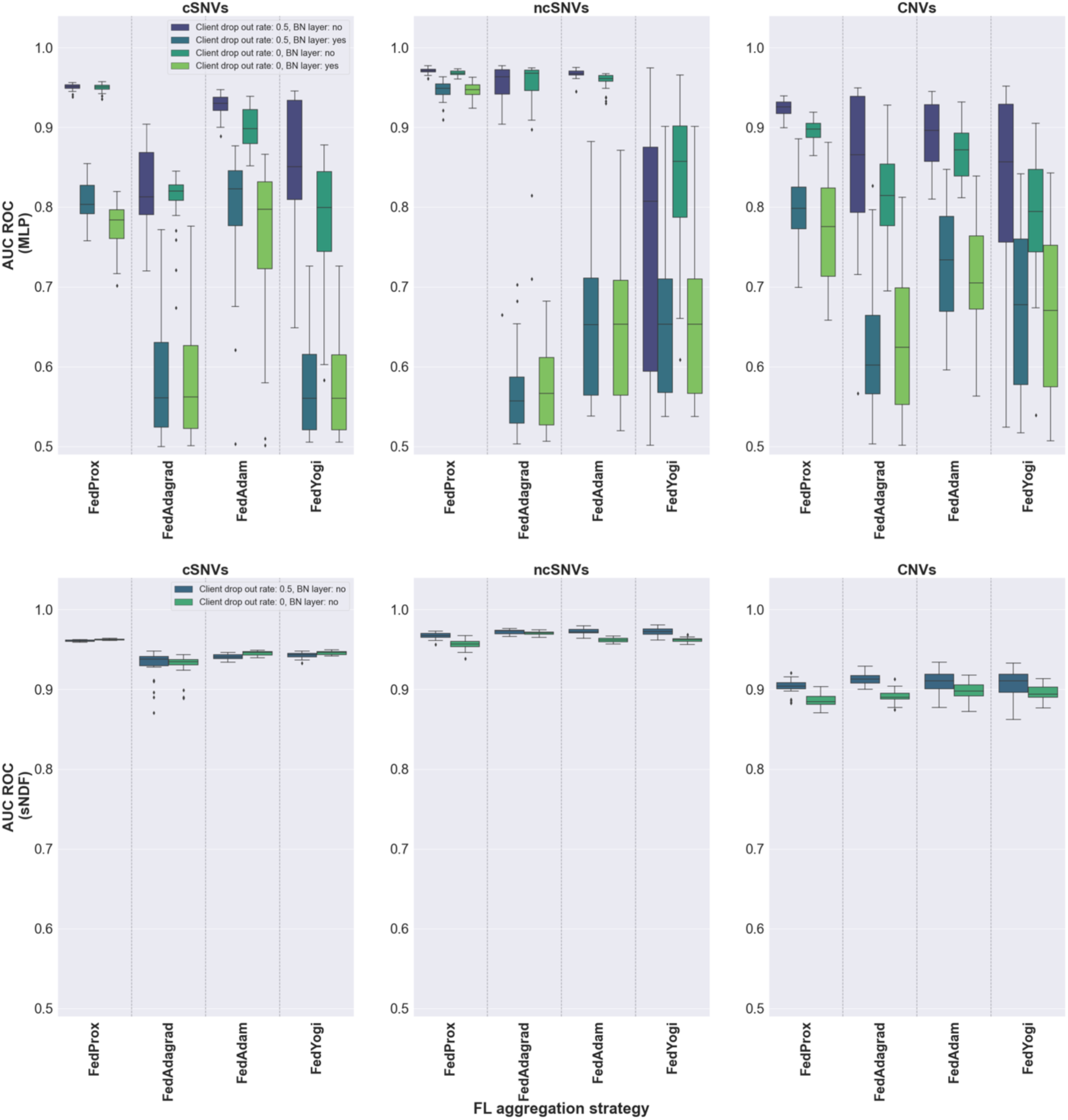
AUC ROC values obtained through cross-validation for different FL aggregation strategies across three types of genetic variants: Coding SNVs (left), non-coding SNVs (middle), and CNVs (right panels). FL aggregation strategies considered are indicated in the x-axis: FedProx, FedAdagrad, FedAdam, and FedYogi. Panels in the first row present the performance of such FL aggregation strategies when applied to the MLP model, with varying FL client participation rates of 50% and 100%, and considering the inclusion or exclusion of local batch normalization layers as indicated in the legend (as shown in the inset in the top-left panel). The second row displays the performance of the FL strategies based on sNDF models, also considering FL client rates of 50% and 100%. Boxplots in the panels represent the distribution of AUC ROC values obtained upon 30 different random seeds for model weight initialization. A perfect classifier would achieve an AUC ROC value equal to 1, while a random classifier would achieve an AUC ROC value of 0.5. Therefore, for all panels, a higher AUC indicates a better performance.

**Supplementary Figure 2.**
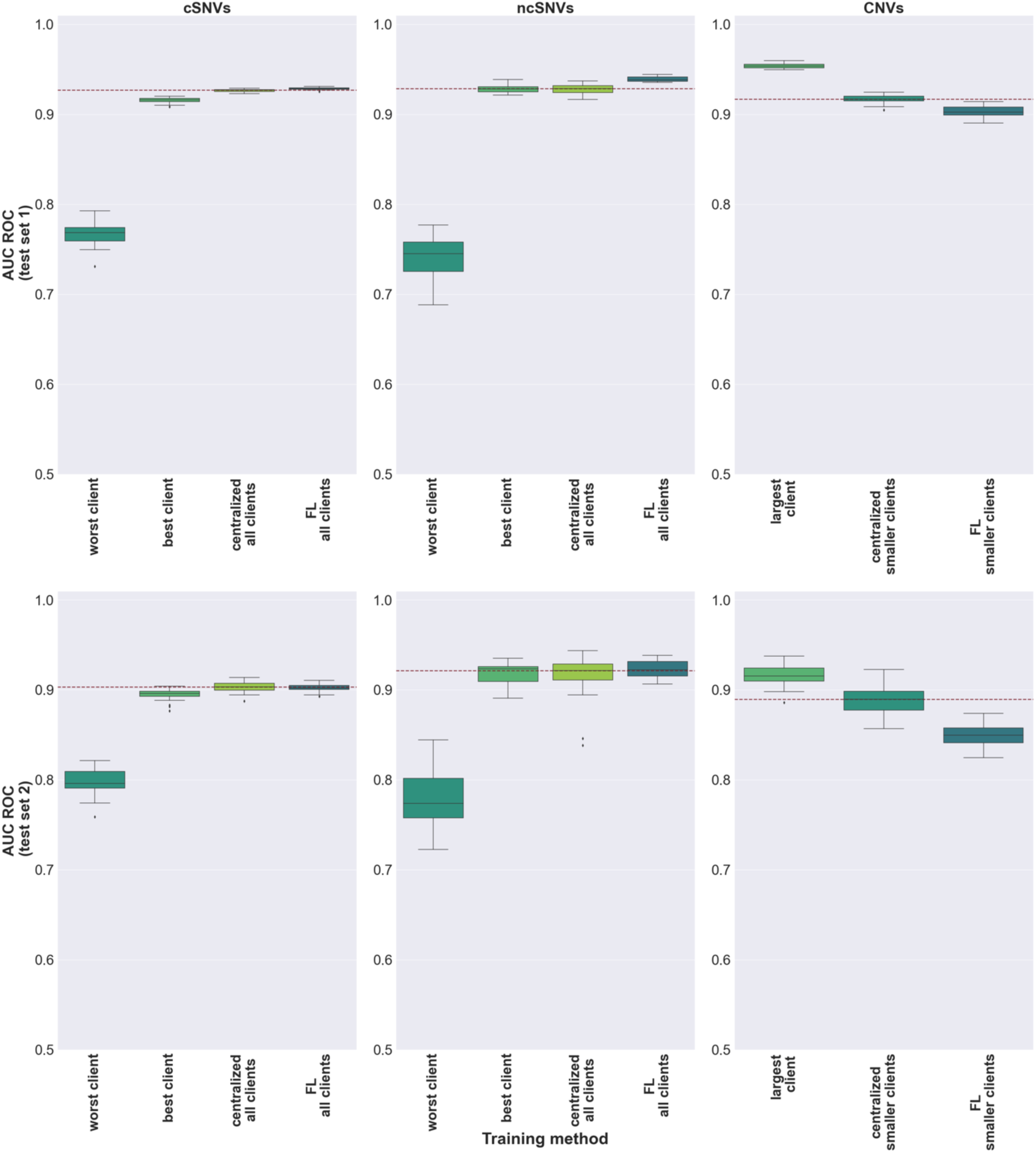
Performance of local, centralized, and federated sNDF models on two independent test sets. Top and bottom panels show, respectively, the results on the first and second independent sets (**Methods**). In the case of coding SNVs and non-coding SNVs, the performance of the worst and best individual client models as well as the pooled-clients models are displayed. The performances of the remaining client models are reported in **Supplementary Tables 7-9**. In the case of CNVs, the performance of the largest client was compared against the centralized and federated learning models of the smaller clients. Boxplots in the panels represent the distribution of AUC ROC values obtained upon 30 different random seeds for model weight initialization. To ease comparison across models, a dotted red line represents the median value obtained for the centralized models.

**Supplementary Figure 3.**
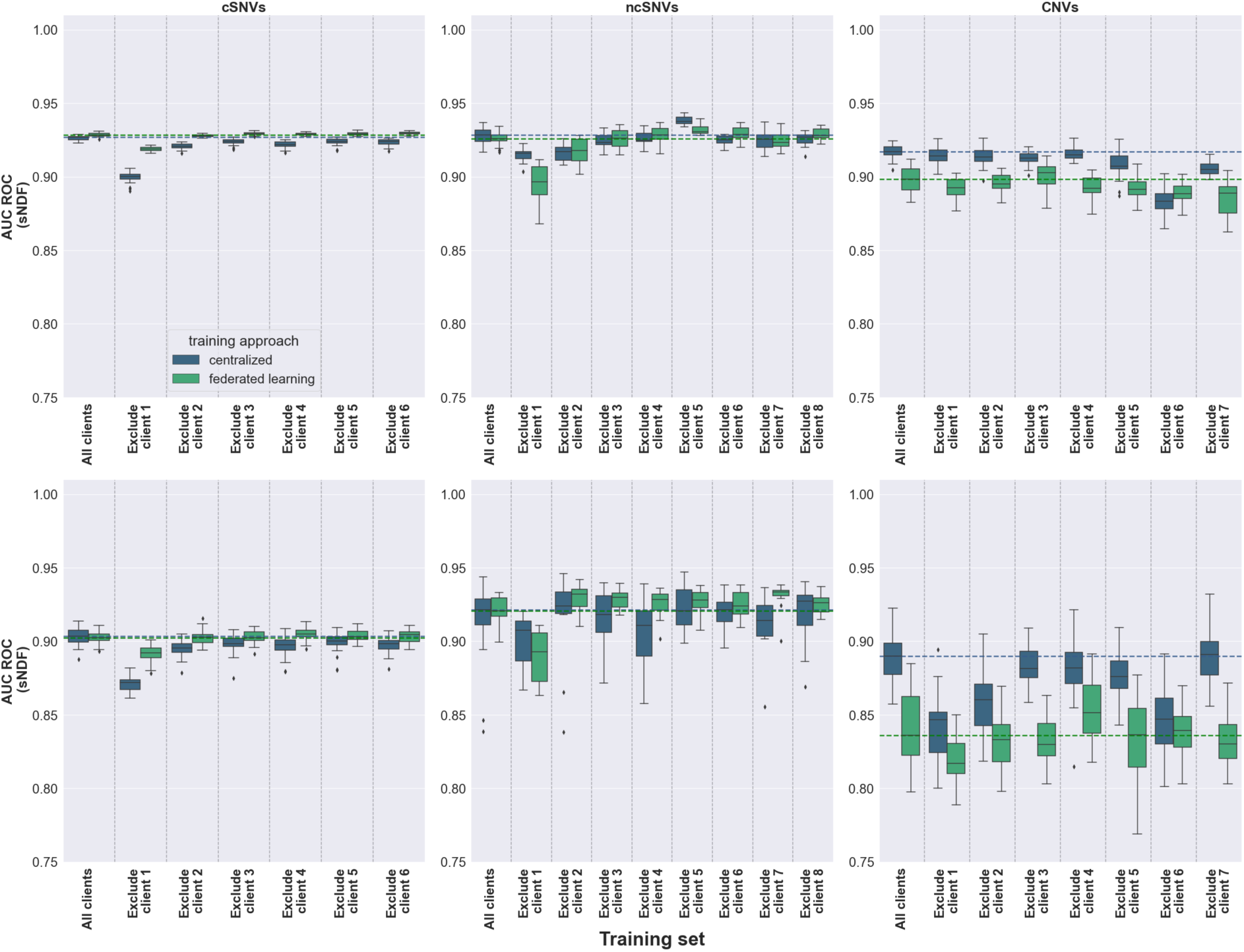
Performance of centralized, and federated sNDF models upon client dropouts on two independent test sets. Top and bottom panels show, respectively, the results obtained on the first and second independent tests (**Methods**). Boxplots in the panels represent the distribution of AUC ROC values obtained upon 30 different random seeds for model weight initialization. Centralized and FL models are colored in blue and green, respectively. Each panel represents the AUC ROC values obtained considering all clients, as well as excluding one client at a time. To ease comparison across models, dotted lines represent the median values obtained for the centralized (blue) and federated (green) models considering all clients.

**Supplementary Figure 4.**
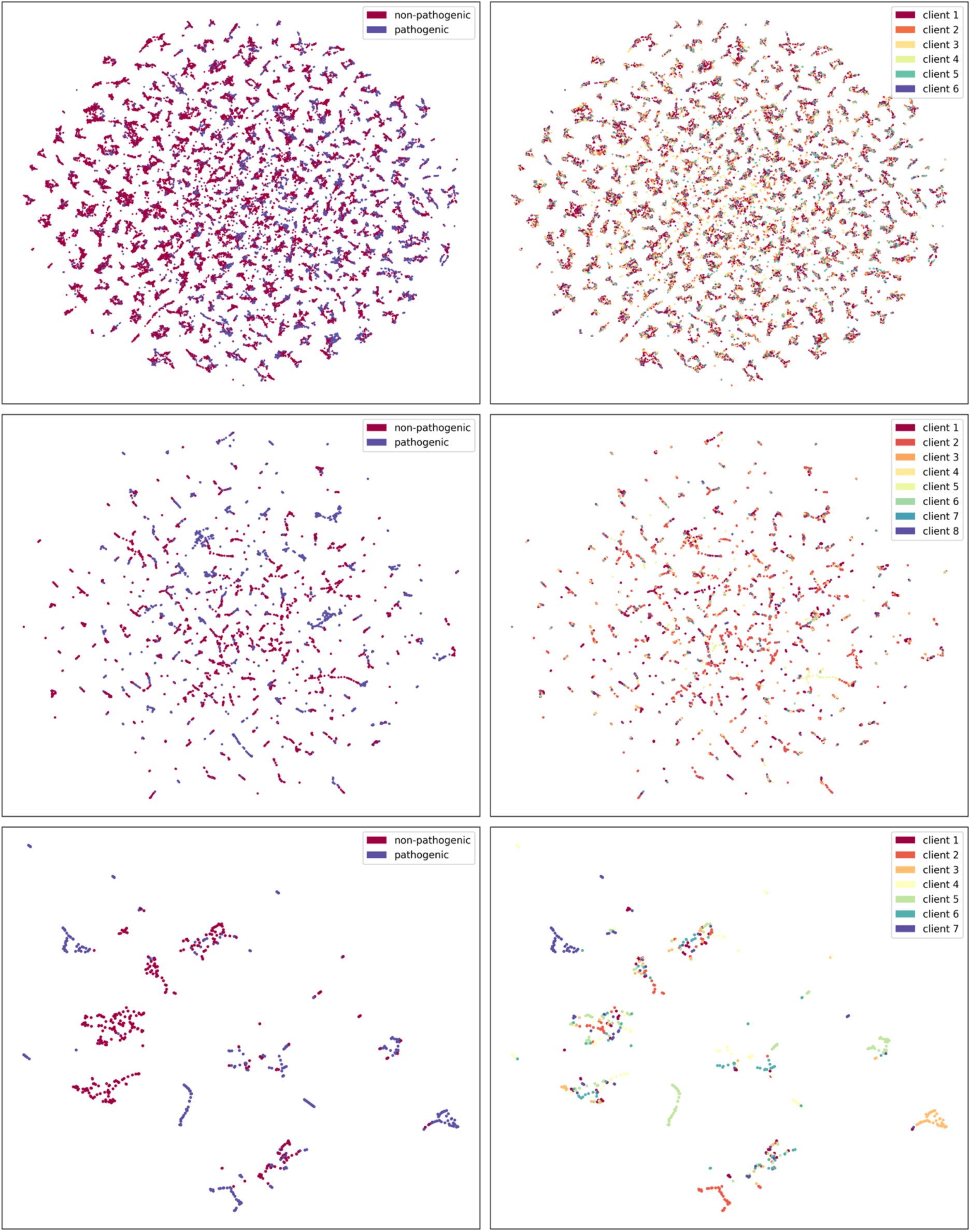
Variant distribution in an unsupervised low-dimensional UMAP representation obtained from the variants feature matrices. Coding SNVs, non-coding SNVs and CNVs are represented in top, middle, and bottom panels, respectively. Panels on the left represent variants colored according to their clinical label: pathogenic (blue) or benign (red). Panels on the right are colored according to their submitter institution (see **Supplementary Tables 2-5**).

### Supplementary Tables

**Supplementary Table 1:** Overview of the collaborative training experiments. The table summarizes the key configurations explored in the collaborative training of machine learning models for the clinical assessment of coding SNVs, non-coding SNVs, and deletion CNVs. It details the number of features used for variant annotation, the models trained, namely Multilayer Perceptron (MLP) and Shallow Neural Decision Forest (sNDF), the baseline models compared for each case study, the federated learning (FL) aggregation strategies employed, and the client participation rates during local training in each round of federated training. Additionally, it outlines the datasets used for evaluation and the evaluation metric applied, which is the area under the ROC curve (AUC ROC).

| Variant type | Coding SNVs | Non-coding SNVs | Deletion CNVs |
| --- | --- | --- | --- |
| Number of features | 60 | 60 | 38 |
| Model comparisons | Single clients vs<br>FL of all clients vs<br>CDS of all clients | Single clients vs<br>FL of all clients vs<br>CDS of all clients | Largest client vs<br>FL of smaller clients vs<br>CDS of smaller clients |
| Learning algorithm | MLP and sNDF |  |  |
| FL aggregation strategy | FedProx, FedAdam, FedAdagrad, FedYogi |  |  |
| Client rate | 50% and 100% |  |  |
| Evaluation | Collaborative Cross-Validation (CCV)<br>Two held-out test sets |  |  |
| Metric | AUC ROC |  |  |

**Supplementary Table 2:** Details of clients in the multi-institutional coding single nucleotide variants dataset. The table provides detailed information about the institutions included in the multi-institutional coding single nucleotide variants dataset derived from the ClinVar database. It lists the names and locations of the contributing institutions, along with the original dataset size and the adjusted size following random under-sampling of the majority class. Additionally, it specifies the number of pathogenic and benign samples associated with each institution. The first column indicates the client id used along the manuscript for this variant type.

| Num-ber id | Institution | Headquarters | Dataset size (original) | Patho-genic | Benign | Dataset size (after random under-sampling) |
| --- | --- | --- | --- | --- | --- | --- |
| 1 | Invitae | San Francisco, California, United States | 30,815 | 8114 | 22,701 | 16,228 |
| 2 | GeneDx | Gaithersburg, Maryland, United States | 6177 | 3945 | 2232 | 4464 |
| 3 | Illumina Laboratory Services, Illumina | San Diego, California, United States | 4680 | 123 | 4557 | 246 |
| 4 | EGL Genetic Diagnostic, Eurofin Clinical Diagnostics | Tucker, Georgia, United States | 1940 | 1143 | 797 | 1594 |
| 5 | Ambry Genetics | Aliso Viejo, California, United States | 2,527 | 1629 | 898 | 1,796 |
| 6 | Laboratory for Molecular Medicine, Partners Healthcare, Personalized Medicine | Boston, Massachusetts, United States | 1417 | 362 | 1055 | 724 |

**Supplementary Table 3:** Details of clients in the multi-institutional non-coding single nucleotide variants dataset. The table provides detailed information about the institutions included in the multi-institutional non-coding single nucleotide variants dataset derived from the ClinVar database. It lists the names and locations of the contributing institutions, along with the original dataset size and the adjusted size following random under-sampling of the majority class. Additionally, it specifies the number of pathogenic and benign samples associated with each institution. The first column indicates the client id used along the manuscript for this variant type.

| Num-<br>ber id | Institution | Headquarters | Dataset<br>size<br>(original) | Patho-<br>genic | Benign | Data size<br>(after<br>random<br>under-sampling) |
| --- | --- | --- | --- | --- | --- | --- |
| 1 | GeneDx | Gaithersburg, Maryland, United States | 2,288 | 560 | 1,728 | 1,120 |
| 2 | Invitae | San Francisco, California, United States | 1,735 | 965 | 770 | 1,540 |
| 3 | EGL Genetic Diagnostics, Eurofins Clinical Diagnostics | Tucker, Georgia, United States | 357 | 288 | 69 | 138 |
| 4 | Laboratory for Molecular Medicine, Partners Healthcare Personalized Medicine | Boston, Massachusetts, United States | 136 | 60 | 76 | 120 |
| 5 | Wong Mito Lab, Molecular and Human Genetics, Baylor College of Medicine | Houston, Texas, United States | 176 | 52 | 124 | 104 |
| 6 | Athena Diagnostics | Marlborough, Massachusetts, United States | 101 | 64 | 37 | 74 |
| 7 | Integrated Genetics/ Laboratory Corporation of America, LabCorp | Westborough, Massachusetts, United States | 108 | 73 | 25 | 50 |
| 8 | ARUP Laboratories, Molecular Genetics and Genomic | Salt Lake City, Utah, United States | 126 | 33 | 93 | 66 |

**Supplementary Table 4:**
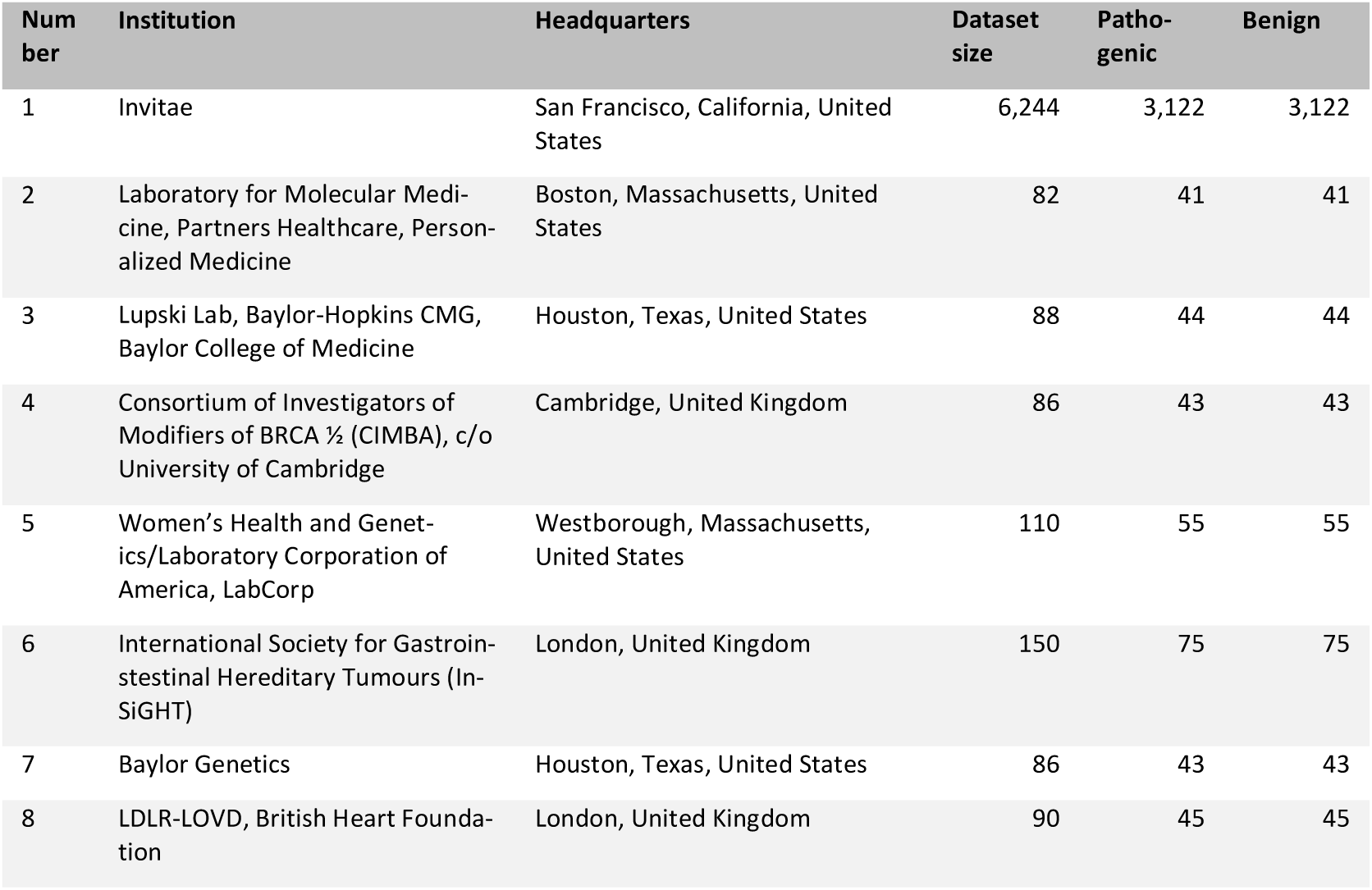
Details of clients in the multi-institutional deletion copy number variants dataset. The table provides detailed information about the institutions included in the multi-institutional deletion copy number variants dataset derived from the ClinVar database. It lists the names and locations of the contributing institutions, along with the original dataset size and the number of pathogenic and benign samples associated with each institution. The first column indicates the client id used along the manuscript for this variant type.

**Supplementary Table 5.** Hyperparameters retained for the centralized and federated models for the pathogenicity annotation of coding SNVs, non-coding SNVs, and CNVs. See Excel file attached.

**Supplementary Table 6:** Size of multi-institutional datasets and independent test sets per genetic variant type. The table summarize the number of institutions, the total number of genetic variants present in the multi-institutional training dataset, and the number of variants in the two independent test sets for each variant type.

| Genetic variant type | No. clients | Training set size | Test set 1 size | Test set 2 size |
| --- | --- | --- | --- | --- |
| Coding SNVs | 6 | 79,556 | 10,378 | 2,838 |
| Non-coding SNVs | 8 | 11,396 | 5,534 | 472 |
| CNVs | 8 | 6,936 | 682 | 96 |

**Supplementary Table 7.**
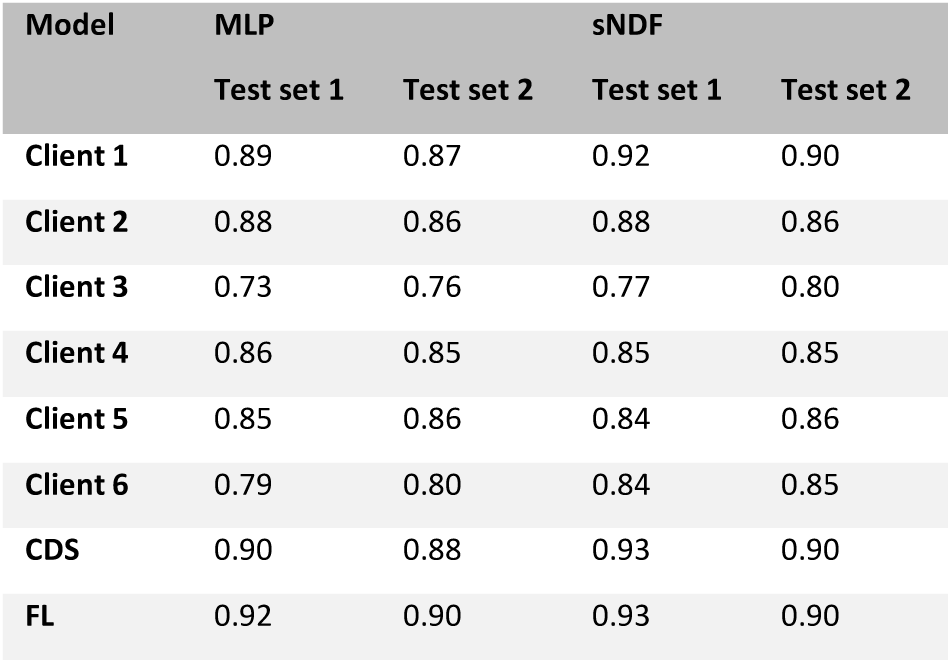
Performance of local, centralized and federated models for the clinical assessment of coding single nucleotide variants. The table presents the mean area under the ROC curve (AUC ROC) values for the clinical assessment of coding single nucleotide variants, comparing local, centralized, and federated models. The performance of the Multilayer Perceptron (MLP) and Shallow Neural Decision Forest (sNDF) models is reported for each approach: local (denoted as client i), centralized (CDS), and federated learning (FL), evaluated across two independent test sets.

**Supplementary Table 8:** Performance of local, centralized and federated models for the clinical assessment of non-coding single nucleotide variants. The table presents the mean area under the ROC curve (AUC) values for the clinical assessment of non-coding single nucleotide variants, comparing local, centralized, and federated models. The performance of the Multilayer Perceptron (MLP) and Shallow Neural Decision Forest (sNDF) models is reported for each approach: local (denoted as client i), centralized (CDS), and federated learning (FL), evaluated across two independent test sets. The results of local model corresponding to client 5 are not listed, since this institution did not have variants found on each chromosome.

| Model | MLP |  | sNDF |  |
| --- | --- | --- | --- | --- |
|  | Test set 1 | Test set 2 | Test set 1 | Test set 2 |
| Client 1 | 0.94 | 0.94 | 0.93 | 0.93 |
| Client 2 | 0.93 | 0.90 | 0.93 | 0.90 |
| Client 3 | 0.85 | 0.86 | 0.86 | 0.87 |
| Client 4 | 0.84 | 0.81 | 0.86 | 0.80 |
| Client 6 | 0.80 | 0.80 | 0.85 | 0.86 |
| Client 7 | 0.69 | 0.70 | 0.75 | 0.78 |
| Client 8 | 0.75 | 0.76 | 0.80 | 0.82 |
| CDS | 0.93 | 0.90 | 0.93 | 0.93 |
| FL | 0.94 | 0.93 | 0.94 | 0.93 |

**Supplementary Table 9:** Performance of largest client, centralized, and federated models of smaller clients for the clinical assessment of deletion copy number variants. The table presents the mean area under the ROC curve (AUC ROC) values for the clinical assessment of deletion copy number variants, comparing the performance of models trained with the largest client, centralized models, and federated models of smaller clients. The table reports the results for the Multilayer Perceptron (MLP) and Shallow Neural Decision Forest (sNDF) models across three approaches: the largest client (Invitae), centralized (CDS), and federated learning (FL), evaluated on two independent test sets.

| Model | MLP |  | sNDF |  |
| --- | --- | --- | --- | --- |
|  | Test set 1 | Test set 2 | Test set 1 | Test set 2 |
| Largest client | 0.96 | 0.92 | 0.96 | 0.92 |
| CDS | 0.90 | 0.87 | 0.92 | 0.89 |
| FL | 0.90 | 0.87 | 0.90 | 0.86 |

**Supplementary Table 10:** Wilcoxon rank-sum test results for centralized and federated models in the clinical assessment of genetic variants. The table presents the results of Wilcoxon rank-sum test, comparing the mean area under the curve ROC obtained by the best-performing centralized and federated learning models across two independent test sets. The Wilcoxon rank-sum statistic and corresponding p-values are reported for each type of genetic variant: coding single nucleotide variant (cSNV), non-coding single nucleotide variant (ncSNV), and deletion copy number variant (CNV). Results are shown for both the Multilayer Perceptron (MLP) and Shallow Neural Decision Forest (sNDF) models.

| Variant type | MLP |  |  |  | sNDF |  |  |  |
| --- | --- | --- | --- | --- | --- | --- | --- | --- |
|  | Test set 1 |  | Test set 2 |  | Test set 1 |  | Test set 2 |  |
|  | Stat. | p-value | Stat. | p-value | Stat. | p-value | Stat. | p-value |
| cSNVs | -6.59 | 4.28e-11 | -6.40 | 1.53e-10 | -5.04 | 4.62e-07 | 0.84 | 0.40 |
| ncSNVs | -5.63 | 1.77e-08 | -4.33 | 1.47e-05 | -6.51 | 7.03e-11 | -0.77 | 0.44 |
| CNVs | -0.99 | 0.32 | -0.62 | 0.53 | 6.28 | 3.31e-10 | 6.20 | 5.57e-10 |

**Supplementary Table 11:** Wilcoxon rank-sum test comparing centralized models before and after dropping a client from the training set. The table presents the results of the Wilcoxon rank-sum tests comparing the mean area under the curve ROC of the best-performing centralized model trained across of all clients and those trained after dropping a given client i. across the two independent test sets. The Wilcoxon rank-sum statistic and corresponding p-values are reported for each type of genetic variant: coding single nucleotide variant (cSNV), non-coding single nucleotide variant (ncSNV), and deletion copy number variant (CNV). Results are shown for both the Multilayer Perceptron (MLP) and Shallow Neural Decision Forest (sNDF) models.

| Variant type | Training set | MLP |  |  |  | sNDF |  |  |  |
| --- | --- | --- | --- | --- | --- | --- | --- | --- | --- |
|  |  | Test set 1 |  | Test set 2 |  | Test set 1 |  | Test set 2 |  |
|  |  | Stat. | p-value | Stat. | p-value | Stat. | p-value | Stat. | p-value |
| cSNVs | Dropping client 1 | 4.54 | 5.66e-06 | 3.81 | 1.36e-04 | 6.65 | 2.87e-11 | 6.65 | 2.87e-11 |
|  | Dropping client 2 | 6.65 | 2.87e-11 | 6.64 | 3.18e-11 | 6.57 | 4.73e-11 | 4.70 | 2.58e-06 |
|  | Dropping client 3 | 6.65 | 2.87e-11 | 6.47 | 9.45e-11 | 4.39 | 1.13e-05 | 3.08 | 2.00e-03 |
|  | Dropping client 4 | 6.65 | 2.87e-11 | 6.53 | 6.37e-11 | 6.23 | 4.40e-10 | 3.70 | 2.19e-04 |
|  | Dropping client 5 | 6.65 | 2.87e-11 | 6.37 | 1.86e-10 | 4.54 | 5.66e-06 | 2.29 | 0.02 |
|  | Dropping client 6 | 6.65 | 2.87e-11 | 6.61 | 3.88e-11 | 4.91 | 9.18e-07 | 3.36 | 7.91e-04 |
| ncSNVs | Dropping client 1 | 6.65 | 2.87e-11 | 6.05 | 1.47e-09 | 4.59 | 4.43e-06 | 2.94 | 3.26e-03 |
|  | Dropping client 2 | 6.65 | 2.87e-11 | 5.81 | 6.24e-09 | 4.32 | 1.52e-05 | -0.62 | 0.54 |
|  | Dropping client 3 | 6.64 | 3.18e-11 | 4.73 | 2.15e-06 | 2.03 | 0.04 | 0.21 | 0.84 |
|  | Dropping client 4 | 6.65 | 2.87e-11 | 6.34 | 2.15e-10 | 0.94 | 0.35 | 1.50 | 0.13 |
|  | Dropping client 5 | 6.65 | 2.87e-11 | 4.95 | 7.31e-07 | -4.71 | 2.50e-06 | -0.19 | 0.85 |
|  | Dropping client 6 | 6.65 | 2.87e-11 | 4.28 | 1.81e-05 | 2.09 | 0.04 | 0.17 | 0.86 |
|  | Dropping client 7 | 6.65 | 2.87e-11 | 5.23 | 1.66e-07 | 1.56 | 0.12 | 1.40 | 0.16 |
|  | Dropping client 8 | 6.65 | 2.87e-11 | 5.23 | 1.66e-07 | 1.59 | 0.11 | -0.22 | 0.83 |
| CNVs | Dropping client 1 | 2.61 | 8.87e-03 | 5.44 | 5.31e-08 | 2.59 | 0.01 | 6.20 | 5.57e-10 |
|  | Dropping client 2 | 0.04 | 0.96 | 1.36 | 0.17 | 2.70 | 0.01 | 5.12 | 3.01e-07 |
|  | Dropping client 3 | 1.43 | 0.15 | -1.52 | 0.13 | 3.78 | 1.58e-04 | 1.42 | 0.16 |
|  | Dropping client 4 | 1.23 | 0.26 | 5.49 | 3.96e-08 | 1.75 | 0.08 | 1.43 | 0.15 |
|  | Dropping client 5 | 1.06 | 0.29 | -1.98 | 0.05 | 4.14 | 3.47e-05 | 2.71 | 0.01 |
|  | Dropping client 6 | 6.26 | 4.01e-10 | 0.21 | 0.83 | 6.65 | 2.87e-11 | 5.97 | 2.43e-09 |
|  | Dropping client 7 | 5.01 | 5.39e-07 | -3.80 | 1.44e-04 | 6.15 | 7.73e-10 | -0.31 | 0.75 |

**Supplementary Table 12:** Wilcoxon rank-sum test comparing federated learning models before and after dropping a client from the training set. This table presents the results of Wilcoxon rank-sum test, comparing the mean area under the curve ROC of the best-performing federated learning model trained across of clients considered initially in the simulations, and after dropping client i. across the two independent test sets. The Wilcoxon rank-sum statistic and corresponding p-values are reported for each type of genetic variant: coding single nucleotide variant (cSNV), non-coding single nucleotide variant (ncSNV), and deletion copy number variant (CNV). Results are shown for both the Multilayer Perceptron (MLP) and Shallow Neural Decision Forest (sNDF) models.

| Variant type | Training set | MLP |  |  |  | sNDF |  |  |  |
| --- | --- | --- | --- | --- | --- | --- | --- | --- | --- |
|  |  | Test set 1 |  | Test set 2 |  | Test set 1 |  | Test set 2 |  |
|  |  | Stat. | p-value | Stat. | p-value | Stat. | p-value | Stat. | p-value |
| cSNVs | Dropping client 1 | 6.21 | 5.32e-10 | 4.92 | 8.51e-07 | 6.65 | 2.87e-11 | 5.91 | 3.34e-09 |
|  | Dropping client 2 | 1.98 | 0.23 | 1.33 | 0.18 | 2.42 | 0.02 | -0.27 | 0.79 |
|  | Dropping client 3 | -0.41 | 0.68 | -0.22 | 0.83 | -2.61 | 0.01 | -0.41 | 0.68 |
|  | Dropping client 4 | 0.38 | 0.70 | -0.17 | 0.86 | -1.82 | 0.07 | -2.94 | 3.25e-03 |
|  | Dropping client 5 | -0.06 | 0.95 | 0.80 | 0.42 | -1.33 | 0.18 | -1.27 | 0.20 |
|  | Dropping client 6 | -0.34 | 0.73 | -0.21 | 0.83 | -3.46 | 5.41e-04 | -1.12 | 0.26 |
| ncSNVs | Dropping client 1 | 5.26 | 1.42e-07 | 1.90 | 0.06 | 4.86 | 1.20e-06 | 4.56 | 5.09e-06 |
|  | Dropping client 2 | 5.92 | 3.06e-09 | 2.76 | 0.01 | 2.82 | 4.73e-03 | -2.47 | 0.01 |
|  | Dropping client 3 | 0.38 | 0.70 | 0.94 | 0.34 | 0.03 | 0.98 | -2.32 | 0.02 |
|  | Dropping client 4 | 0.04 | 0.96 | -0.93 | 0.35 | -1.29 | 0.19 | -1.50 | 0.13 |
|  | Dropping client 5 | -0.96 | 0.34 | -0.87 | 0.38 | -3.53 | 4.14e-04 | -2.04 | 0.04 |
|  | Dropping client 6 | 0.89 | 0.37 | -0.45 | 0.65 | -1.85 | 0.06 | -1.27 | 0.21 |
|  | Dropping client 7 | -0.80 | 0.42 | -0.66 | 0.51 | 1.26 | 0.21 | -3.41 | 6.41e-04 |
|  | Dropping client 8 | 0.24 | 0.81 | -1.29 | 0.20 | -1.70 | 0.09 | -1.06 | 0.29 |
| CNVs | Dropping client 1 | 0.50 | 0.62 | 2.65 | 8.14e-03 | 2.74 | 0.01 | 3.49 | 4.71e-04 |
|  | Dropping client 2 | -0.81 | 0.42 | -0.55 | 0.58 | 1.28 | 0.20 | 1.22 | 0.21 |
|  | Dropping client 3 | -3.56 | 3.67e-04 | 0.62 | 0.53 | -1.47 | 0.13 | 1.25 | 0.21 |
|  | Dropping client 4 | 1.29 | 0.20 | 0.59 | 0.55 | 2.23 | 0.02 | -2.29 | 0.02 |
|  | Dropping client 5 | -0.89 | 0.38 | 1.18 | 0.24 | 2.96 | 3.11e-03 | 0.98 | 0.33 |
|  | Dropping client 6 | 3.58 | 3.46e-04 | 1.79 | 0.07 | 3.70 | 2.18e-04 | 0.03 | 0.97 |
|  | Dropping client 7 | 1.11 | 0.27 | -3.17 | 1.51 | 4.05 | 5.10e-05 | 1.25 | 0.21 |

**Supplementary Table 13.** Spearman’s rank correlation test results for centralized and federated models in the clinical assessment of genetic variants. The table presents the results of Spearman’s rank correlation test, comparing the pathogenicity predictions of the best-performing centralized and federated learning models across two independent test sets. The Spearman’s rank correlation coefficient and the associated p-value are reported for each genetic variant type: coding single nucleotide variants (cSNVs), non-coding single nucleotide variants (ncSNVs), and deletion copy number variants (CNVs). Results are shown for both the Multilayer Perceptron (MLP) and Shallow Neural Decision Forest (sNDF) models.

| Variant type | MLP |  |  |  | sNDF |  |  |  |
| --- | --- | --- | --- | --- | --- | --- | --- | --- |
|  | Test set 1 |  | Test set 2 |  | Test set 1 |  | Test set 2 |  |
|  | Stat. | p-value | Stat. | p-value | Stat. | p-value | Stat. | p-value |
| cSNVs | 0.93 | 6.03e-303 | 0.94 | 6.88e-298 | 0.95 | 2.16e-132 | 0.96 | 6.45e-307 |
| ncSNVs | 0.93 | 1.78e-277 | 0.94 | 3.26e-228 | 0.94 | 4.62e-145 | 0.97 | 4.78e-291 |
| CNVs | 0.88 | 3.32e-224 | 0.86 | 1.89e-30 | 0.87 | 4.53e-213 | 0.82 | 1.36e-25 |

## List of abbreviations

AUC ROC: Area Under the Receiver Operating Characteristic curve
CDS: Collaborative Data Sharing
CCR: Constrained Coding Region
CNN: Convolutional Neural Network
CNV: Copy Number Variant
FL: Federated Learning
GDPR: General Data Protection Regulation
GWAS: Genome-Wide Association Studies
HARs: Human Accelerated Regions
HPC: High-Performance Computing
IID: Independent and Identically Distributed
LADs: Lamina-Associated Domains
MLP: Multilayer Perceptron
ML: Machine Learning
sNDF: Shallow Neural Decision Forest
SNV: Single-Nucleotide Variant
SV: Structural Variant
TSS: Transcription Start Site
TPM: Transcripts Per Million
UCNEs: Ultra-Conserved Non-Coding Elements
UMAP: Uniform Manifold Approximation and Projection
WGS: Whole Genome Sequencing

